# Antibody binding and ACE2 binding inhibition is significantly reduced for the Omicron variant compared to all other variants of concern

**DOI:** 10.1101/2021.12.30.21267519

**Authors:** Daniel Junker, Matthias Becker, Teresa R. Wagner, Philipp D. Kaiser, Sandra Maier, Tanja M. Grimm, Johanna Griesbaum, Patrick Marsall, Jens Gruber, Bjoern Traenkle, Constanze Heinzel, Yudi T. Pinilla, Jana Held, Rolf Fendel, Andrea Kreidenweiss, Annika Nelde, Yacine Maringer, Sarah Schroeder, Juliane S. Walz, Karina Althaus, Gunalp Uzun, Marco Mikus, Tamam Bakchoul, Katja Schenke-Layland, Stefanie Bunk, Helene Haeberle, Siri Göpel, Michael Bitzer, Hanna Renk, Jonathan Remppis, Corinna Engel, Axel R. Franz, Manuela Harries, Barbora Kessel, Monika Strengert, Gerard Krause, Anne Zeck, Ulrich Rothbauer, Alex Dulovic, Nicole Schneiderhan-Marra

## Abstract

The rapid emergence of the Omicron variant and its large number of mutations has led to its classification as a variant of concern (VOC) by the WHO(1). Initial studies on the neutralizing response towards this variant within convalescent and vaccinated individuals have identified substantial reductions(2-8). However many of these sample sets used in these studies were either small, uniform in nature, or were compared only to wild-type (WT) or, at most, a few other VOC. Here, we assessed IgG binding, (Angiotensin-Converting Enzyme 2) ACE2 binding inhibition, and antibody binding dynamics for the omicron variant compared to all other VOC and variants of interest (VOI)(9), in a large cohort of infected, vaccinated, and infected and then vaccinated individuals. While omicron was capable of binding to ACE2 efficiently, antibodies elicited by infection or immunization showed reduced IgG binding and ACE2 binding inhibition compared to WT and all VOC. Among vaccinated samples, antibody binding responses towards omicron were only improved following administration of a third dose. Overall, our results identify that omicron can still bind ACE2 while pre-existing antibodies can bind omicron. The extent of the mutations appear to inhibit the development of a neutralizing response, and as a result, omicron remains capable of evading immune control.

## Main Text

Since its initial outbreak in late 2019, SARS-CoV-2 has morphed into a global pandemic, characterised by waves of infection within countries and regions. Following the initial global wave, subsequent waves have often been triggered by the emergence of variants of concern that outcompeted earlier variants, which have increased transmission or show the ability to escape vaccine and infection-derived immunity (10-15). Several of these variants have been classified as variants of concern (VOC) by the WHO (9), with the delta variant currently being responsible for the majority of active infections globally (16). Due to concerns that its numerous spike protein mutations would render it able to escape immune control and its rapid spread in South Africa, the WHO classified omicron as a VOC on the 26^th^ of November 2021 (1). Within days of this classification, omicron had already been reported in multiple other countries and has now been shown to be responsible for increasing case numbers in South Africa(17), while also appearing to have a higher risk of reinfection(18). Early studies into neutralization against omicron have identified significant reductions in neutralizing activity, especially for those who have not received three vaccine doses (2-4, 19). However these studies have also often featured a low sample number, a limited diversity of sample types (e.g. only those vaccinated with Pfizer BNT162b2), or directly compared omicron to WT or a single other variant of concern. We provide here a comprehensive analysis of the binding capacity, binding dynamics and ACE2 binding inhibition of omicron towards antibodies generated through either vaccination or natural infection compared to WT and all other VOCs as well as the lambda and mu variants of interest (VOI). In contrast to these studies, we have included samples from a range of vaccines and dosing schemes currently available within the EU including boosters, and from infected samples from both adults and children from the first, second and third waves in Germany.

We analyzed antibody binding responses towards omicron, all other variants of concern/interest and wild-type in sera from individuals with either vaccine-induced (vaccinated), naturally-induced (convalescent) or naturally-induced and vaccine-boosted immunity (infected and vaccinated). We included samples from convalescent individuals from all three waves in Germany so far (first wave, WT n=50, second wave, alpha n=30, third wave, delta n=6), individuals vaccinated twice with two homologous doses of either mRNA-1273 (n=46), AZD1222 (n=60) or BNT162b2 (n=60), individuals vaccinated twice with two heterologous doses of either AZD1222 and mRNA-1273 (n=20) or BNT162b2 (n=20), individuals vaccinated three times with BNT162b2 (n=20) or individuals with a previous SARS-CoV-2 infection who were later vaccinated with a single dose of BNT162b2 (n=25). As controls for unvaccinated individuals, commercially available pre-pandemic sera were used (n=15). Please consult Table 1 for a full description of the characteristics of the samples used in this study.

**Table 1.** Overview of sample characteristics for our study population.

| Sample type | Sub-group | n | Median dT in days (IQR) | Number of females (%) | Median age in years (IQR) | History of immuno-suppressive condition or medication (%) |
| --- | --- | --- | --- | --- | --- | --- |
| Infected | WT (adults) | 30 | 104 (94-119) | 14 (47) | 62 (51-69) | 0 (0) |
|  | WT (children) | 20 | 124 (116-129) | 7 (35) | 11 (7-14) | 0 (0) |
|  | Alpha | 30 | 88 (47-104) | 12 (40) | 56 (42-65) | 14 (47) |
|  | Delta | 6 | 18 (10-23) | 5 (83) | 65 (56-73) | 4 (67) |
| Infected and vaccinated |  | 25 | 54 (23-91) | 16 (64) | 55 (48-59) | 1 (4) |
| Vaccinated | A/A (1-2 mo.) | 30 | 49 (48-52) | 20 (67) | 64 (60-66) | 2 (7) |
|  | A/A (4-6 mo.) | 30 | 154 (146-158) | 23 (77) | 55 (48-60) | 0 (0) |
|  | M/M (1-2 mo.) | 30 | 51 (48-54) | 20 (67) | 59 (49-61) | 1 (3) |
|  | M/M (4-6 mo.) | 16 | 139 (131-145) | 9 (56) | 70 (51-83) | 1 (6) |
|  | P/P (1-2 mo.) | 30 | 51 (49-54) | 20 (67) | 58 (52-66) | 1 (3) |
|  | P/P (4-6 mo.) | 30 | 152 (141-160) | 25 (83) | 38 (30-53) | 0 (0) |
|  | A/M | 20 | 153 (150-154) | 16 (80) | 41 (29-56) | 0 (0) |
|  | A/P | 20 | 151 (144-157) | 19 (95) | 48 (42-56) | 0 (0) |
|  | P/P/P | 20 | 14 (14-26.5) | 13 (65) | 33 (29-44) | 2 (12)* |
| Negative |  | 15 | - | 8 (53) | 37 (29-41) | 0 (0) |

WT = wild-type (B.1 isolate). A/A = two-dose AZD1222. M/M = two-dose mRNA-1273. P/P = two-dose BNT162b2. A/M = first dose AZD1222, second dose mRNA-1273. A/P = first dose AZD1222, second dose BNT162b2. P/P/P = three-dose BNT162b2. n= number of samples. IQR = interquartile range.

Initially, we examined IgG binding using MULTICOV-AB(20), a previously published SARS-CoV-2 multiplex immunoassay that was adapted to analyze binding towards RBDs from all VOC/VOI (a full list of antigens analyzed and their mutations contained within can be found as Table 2). For both vaccinated and convalescent samples, IgG binding towards omicron from pre-existing antibodies was significantly reduced compared to WT (vaccinated 4.5x fold decrease, infected 8.6 fold, Supplementary Table 1, Figure 1) and all other VOC/VOI (Figure 1a, Supplementary Table 2). By comparison, beta and mu which had the next largest decreases in IgG binding were only reduced 2.6-3.2 fold and 2.9-3.4 fold respectively (Supplementary Table 1).

**Table 2.** Overview of antigens used in MULTICOV-AB and RBDCoV-ACE2.

| Antigen | Manufacturer | Category Number | Mutations covered |
| --- | --- | --- | --- |
| Spike wild-type (B.1) | NMI | - | - |
| RBD wild-type (B.1) | NMI | - | - |
| S1 domain wild-type (B.1) | NMI | - | - |
| S2 domain wild-type (B.1) | Sino Biological | 40590-V08B | - |
| Nucleocapsid wild-type (B.1) | Aalto Bioreagents | 6404-b | - |
| RBD alpha (B.1.1.7) | NMI | - | N501Y |
| RBD beta (B.1.351) | NMI | - | K417N, E484K, N501Y |
| RBD gamma (P1) | NMI | - | K417T, E484K, N501Y |
| RBD delta (B.1.617.2) | NMI | - | L452R, T478K |
| RBD omicron (B.1.529) | Sino Biological | 40592-V08H121 | G339D, S371L, S373P, S375F, K417N, N440K, G446S, S477N, T478K, E484A, Q493R, G496S, Q498R, N501Y, Y505H |
| Spike omicron (B.1.529) | Sino Biological | 40589-V08H26 | A67V, del HV69/70, T95I, G142D, del VYY 143-145, del N211, L212I, ins214EPE, G339D, S371L, S373P, S375F, K417N, N440K, G446S, S477N, T478K, E484A, Q493R, G496S, Q498R, N501Y, Y505H, T547K, H655Y, N679K, P681H, N764K, D796Y, F817P, N856K, A892P, A899P, A942P, Q954H, N969K, L981F, K986P, V987P |
| RBD lambda (C.37) | NMI | - | L452Q, F490S |
| RBD mu (B.1.621) | NMI | - | R346K, E484K, N501Y |

Mutations present within each antigen are provided. Where appropriate, the manufacturers category number is provided. For details on the NMI antigen production, please consult (20, 21, 25).

**Figure 1.**
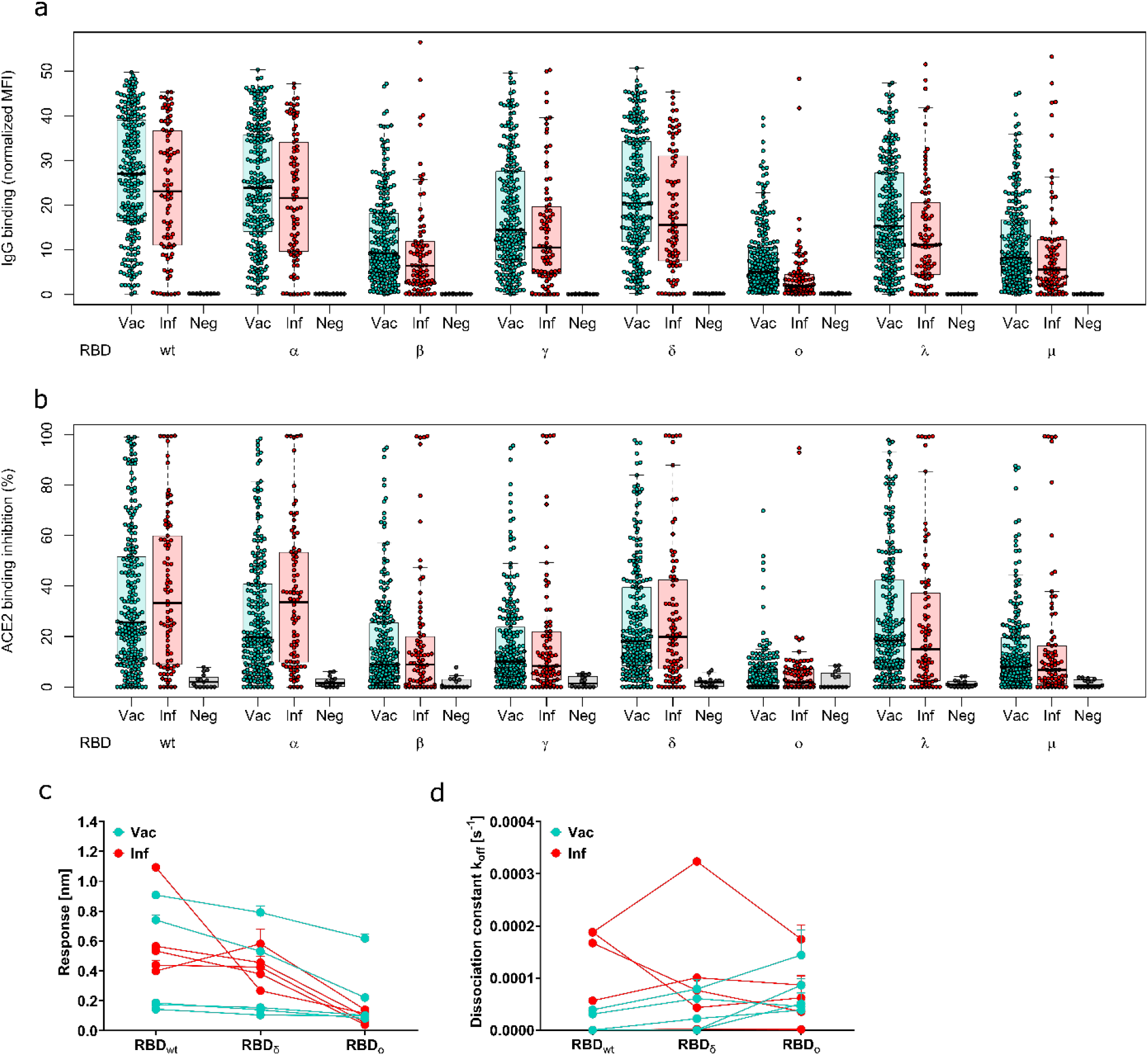
Antibody binding response is significantly reduced for Omicron compared to all other variants of concern. Binding response by pre-existing antibodies generated through either infection or vaccination was measured with MULTICOV-AB (a), RBDCoV-ACE2 (b) and Biolayer interferometry (c and d). (a) Boxplot showing that IgG binding is significantly reduced for omicron compared to all other variants of concern for both infected (n=86) and vaccinated samples (n=226). Negative samples are included as controls (n=15). (b) Boxplot showing that ACE2 binding inhibition is significantly reduced for Omicron compared to all other variants of concern for both infected and vaccinated samples. Boxes represent the median, 25^th^ and 75^th^ percentiles, whiskers show the largest and smallest non-outlier values. Outliers were determined by 1.5 IQR. (c and d) Binding kinetics of RBD specific antibodies from serum samples of convalescent and vaccinated individuals (both n=5). Binding response (c) and dissociation constant (d) were determined by 1:1 fitting model of the individual serum samples between the different RBD variants.

Delta which currently still comprises the majority of global infections was reduced 1.2-1.3 fold compared to WT, and thus was still 3.5-6.3 fold greater than omicron (Supplementary Table 1 and 2). Next, we analyzed ACE2 binding inhibition as proxy for neutralization using RBDCoV-ACE2(21), a multiplex ACE2-RBD inhibition assay. This assay mimics the interaction between ACE2 and the RBD, analyzing the presence of antibodies which block said interaction, while the multiplex format allows for the simultaneous measurement of all VOCs/VUIs in a single well. To evaluate binding against other antigens of SARS-CoV-2, we also include the Spike and S1 domain of WT and the Spike of omicron. In contrast to IgG binding, ACE2 binding inhibition against the omicron RBD was substantially reduced compared to WT (omicron median 0-1.9%, WT median 25.6-33.3%), with only 4.5% of samples considered responsive (Figure 1b, Supplementary Table 3). By contrast, ACE2 binding inhibition for beta, gamma and mu which had the largest decreases relative to WT (median 8.8-8.9%, median 8.3-10.0% and median 6.8-8.0% respectively), were still significantly more responsive compared to omicron. To confirm this lack of RBD binding, we analyzed the binding kinetics of RBD-specific antibodies from vaccinated (two dose BNT162b2) and convalescent (WT) study participants by biolayer interferometry (BLI) analysis (Figure 1c and d). Binding response and dissociation constant were measured for each sample as an indicator of amount and binding strength. Binding response towards the omicron RBD was reduced compared to both WT and delta (Figure 1c), while dissociation increased from WT to omicron for most samples (Figure 1d). While there was a large degree of variation among both the vaccinated and infected samples, omicron always had the lowest binding response (Supplementary Figure 1, Supplementary Table 3). When binding towards ACE2 itself was examined, omicron was still able to bind ACE2 with high affinity (Supplementary Figure 2).

As all other publications on neutralization against omicron had involved either pseudoviruses expressing the full-length spike (2, 5-7), or the native virus itself (3, 8, 19), we analyzed omicron ACE2 binding inhibition against the Spike protein. Omicron ACE2 binding inhibition towards the Spike protein, while still significantly reduced compared to WT (p<0.001), was still present in 6.8% of samples (4.5% for RBD) (Figure 2a). In contrast, IgG binding capacity towards the Spike was more conserved than the RBD (Figure 2 b-d).

**Figure 2.**
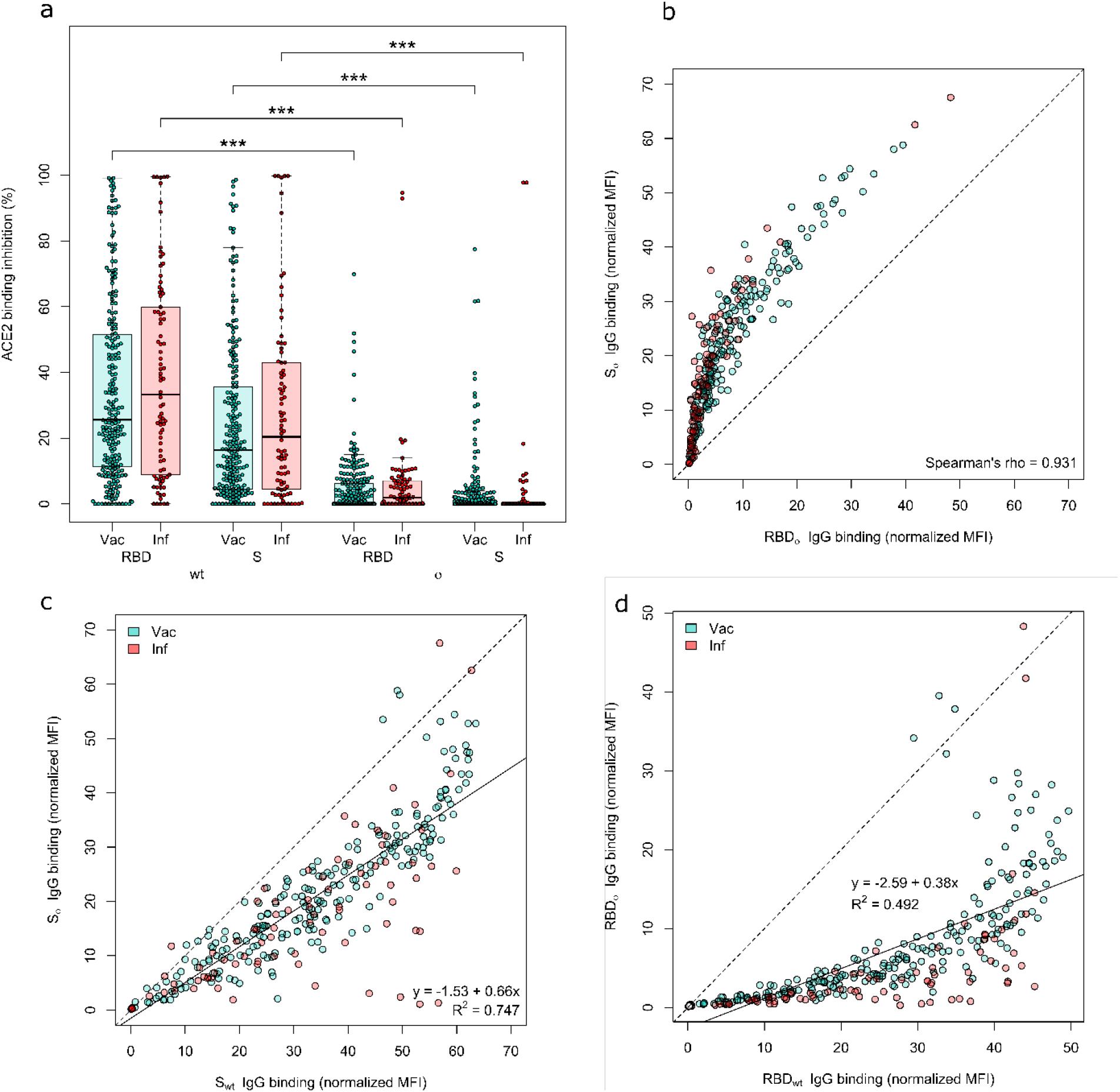
Both the Omicron RBD and Spike protein have minimal neutralizing activity. ACE2 binding inhibition (a) and IgG binding capacity (b-d) were compared for the Omicron RBD and S to the wild-type RBD and S. (a) Boxplot showing that ACE2 binding inhibition while significantly reduced for Omicron in general, is significantly reduced for the RBD compared to the Spike for both Vaccinated (n-=226) and infected (n=86) samples. Boxes represent the median, 25^th^ and 75^th^ percentiles, whiskers show the largest and smallest non-outlier values. Outliers were determined by 1.5 IQR. (b) Correlation analysis of IgG binding capacity for the Omicron spike compared to the Omicron RBD. Spearman’s rank was calculated to assess ordinal association between the variables. (c) Linear regression of IgG binding capacity for the Omicron Spike compared to wild-type Spike. (d) Linear regression of IgG binding capacity for the Omicron RBD compared to wild-type RBD. Statistical significance was calculated by Mann-Whitney U with *** indicating a p-value <0.001.

Next, we analyzed whether vaccine type (AZD1222, mRNA-1273 or BNT162b2) and number of doses (homologous or heterologous 2 dose, or homologous 3 dose) received resulted in differences with respect to omicron binding response. To analyze how this response changed as time post-vaccination increased, we also compared the responses at 1-2 months post-second dose and 5-6 months post-second dose for the homologous recipients. IgG binding capacity at 5-6 months was low for all recipients regardless of the vaccine received, although homologous vector-based vaccination still resulted in lower binding responses than either mRNA-based or heterologous vaccination (Figure 3a). Among samples from 1-2 months post-dosing, infected and then vaccinated had the greatest IgG binding response (median=19.7), followed by two-dose mRNA-1273 (median=12.8), BNT162b2 (median=8.1) and AZD1222 (median=2.3). This pattern of decreasing responses as time post-dose increases remains consistent for ACE2 binding inhibition (Figure 3b). To determine whether a third dose results in increased binding responses against omicron, we analyzed samples from individuals who had received a third dose of BNT162b2 and compared it to those who had received their second dose in a similar timeframe, and individuals who would be eligible to now receive a third dose (Figure 3c, Supplementary Figures 3 and 4). Further boosting was associated with higher omicron ACE2 binding inhibition compared to two doses, suggesting that boosting offers increased protection against omicron. However, this increase in protection was not limited to omicron and was present for all VOC (Figure 3c). Compared to the second dose from a similar timepoint, boosting with the third dose increased substantially confirming this effect is generated by the third dose itself, and not by time-post vaccination alone (Supplementary Figure 3 and 4).

**Figure 3.**
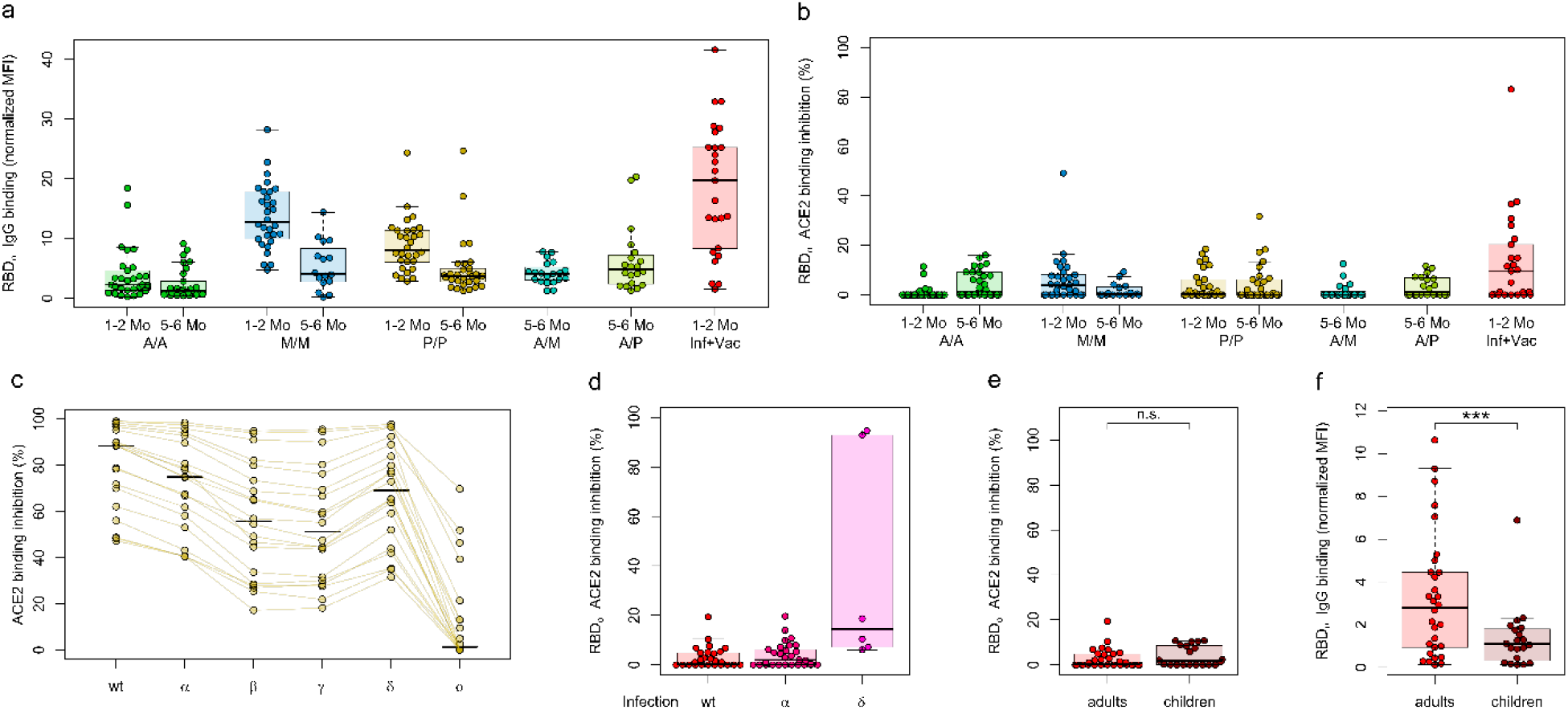
Differences in Omicron binding response among different populations of vaccinated and infected samples. Binding response towards Omicron was analyzed by either MULTICOV-AB (b,f) or RBDCoV-ACE2 (a,c – e). (a and b) Differences in IgG binding and ACE2 binding inhibition by different vaccine schemes (n=30 for samples except for mRNA-1273 5-6 months (n=16), heterologous vaccine schemes (both n=20) and infected and vaccinated (n=25). To determine the effect of time post-vaccination, samples from both 1-2 months and 5-6 months post-vaccination were included. (c) Changes in ACE2 binding response following the third dose of BNT162b2 for all variants of concern and wild-type (n=20). For comparison, both 1-2 months and 5-6 months post second dose BNT162b2 can be found as Supplementary figure 3 and 4 (d) There are no differences in ACE2 binding inhibition towards Omicron for individuals infected with WT (n=30), alpha (n=30) or delta (n=30). (e) Children (n=20) have similar ACE2 binding inhibition towards Omicron compared to adults (n=30) following a WT infection, while showing reduced IgG binding capacity (f). For panels a, b and d-f, boxes represent the median, 25^th^ and 75^th^ percentiles, whiskers show the largest and smallest non-outlier values. Outliers were determined by 1.5 IQR. For panel c, individual samples are highlighted by connected lines with bars representing medians. Statistical significance was calculated by Mann-Whitney U were for e and f with n.s. indicating a p-value <0.05 and *** indicating a p-value<0.001.

Lastly, we analyzed whether natural infection with different variants resulted in differences in binding responses. There was no difference in ACE2 binding inhibition between convalescent individuals infected with either WT or alpha, with both having (Figure 3d) minimal inhibition against omicron. While some samples with a previous delta infection showed substantially more activity compared to WT or alpha, they had been collected much sooner after the infection (median dT 18 days) than WT (median dT 104 days) or alpha (median dT 88 days). To evaluate whether children’s antibodies were more effective at binding towards omicron than adults, we compared convalescent samples 3-4 months post-PCR from the first wave in children (n=20), to convalescent samples 3 months post-positive PCR from the same wave in adults (n=30) (Figure 3e and f). There was no difference between adults and children in ACE2 binding inhibition (Figure 3e), although children did have significantly reduced binding capacity towards omicron (p<0.001) (Figure 3f).

In this study, we provide an in-depth characterization of antibody binding to omicron as compared to WT and all other VOCs and VUIs in a large diverse sample cohort. The use of an ACE2 inhibition assay enabled the comparison of multiple variants of interest simultaneously and was neutralizing antibody specific, while also producing comparable results to classical virus neutralization assays(21). Antibody binding towards the omicron variant elicited by either immunization or previous infection, is substantially reduced when compared with other variants.

Our data are in line with several other initial reports (2-8, 22) and provide additional information on IgG binding capacity and ACE2 binding inhibition by the inclusion of a large variety of sub-cohorts representing the present diversity of immunity against SARS-CoV-2, and by the comparison of the omicron variant to all other VOC/VOI. We found that both IgG binding capacity and ACE2 binding inhibition was more severely reduced for all samples against the omicron variant, than against any other VOC/VOI, with most samples being classified as non-responsive towards omicron for ACE2 binding inhibition. Like others (4, 5, 7), we found that antibody binding responses towards omicron were significantly increased upon administration of booster dose. Boosting resulted in increases in both IgG binding capacity and ACE2 binding inhibition against all VOCs which is important given that delta currently remains the predominant global variant. However, caution should be applied to these results as it remains unknown if this increase in protection following the third dose changes as time post-administration increases. This increased protection following boosting also appears to apply for convalescent individuals, as seen by the increased IgG binding capacity and ACE2 binding inhibition for previously infected individuals who received a single dose, compared to those who had received any two vaccine doses. This increase in responses for individuals who have been both infected and vaccinated is in agreement with (23), who found that infections prior to vaccination resulted in a greater breadth of immune response, while (6) found that breakthrough delta infections among vaccinated individuals acted like a booster dose. Thus both reinfection and a booster dose leads to appropriate affinity maturation of elicited antibodies.

Our results are in agreement that either vaccine protection from two doses or natural immunity generated by a previous infection, appears broadly ineffective against omicron (18). Among those who had received two doses, binding responses were consistent with other reports (2) in identifying a significant decrease for those who received homologous two-dose vector vaccination as opposed to homologous mRNA vaccination or heterologous vaccination.

We identified no difference in binding response for children compared to adults. While previous research has identified that children’s antibody titres are higher than adults following infection (24), this did not result in an increase in binding capacity or ACE2 binding inhibition toward omicron. However, a limitation of our study is that the children and adult groups were not well-matched in terms of time post-positive PCR (median dT 104 for adults, 124 for children) or disease severity (majority hospitalized adults versus asymptomatic/light symptoms children). A larger investigation including vaccinated samples from children is needed to investigate any possible protective effect from previous infection and the antibody response towards Omicron itself in children in more detail.

Similarly to (22), we identified the same pattern of a much less pronounced reduction in RBD IgG binding response compared to RBD-directed neutralizing activity. Our analysis of both the Spike and RBD derived ACE2 binding inhibition suggests that this is consistent for both. Given that the Spike-derived antibodies had the same pattern of decreased neutralizing activity compared to IgG binding response, this suggests that while both epitopes are sufficiently conserved to enable binding, their divergent mutated sequences affect their neutralizing response. Further investigation into this pattern and particularly the role of S1-derived antibodies in neutralization is required to understand the neutralizing protection offered against omicron.

Overall, our results identify that while omicron can still can bind ACE2 and pre-existing antibodies can bind omicron, the extent of the mutations within the RBD appear to be too divergent to enable RBD-directed antibodies to mount a neutralizing response. While antibodies against the Spike protein have increased IgG binding capacity, they also have a significantly reduced neutralizing activity. The dramatic reductions in both IgG binding and ACE2 binding inhibition towards omicron as opposed to other VOC/VOI, confirm this variant remains capable of immune escape and requires careful sequence monitoring to identify any further sequence evolution. Importantly, booster doses elicit a significant increase in antibody response which correlates with a significant increase in both IgG binding and ACE2 binding inhibition against omicron. Our data adds weight a growing body of evidence that the continuous adaptation of vaccines to novel highly contagious variants needs to be considered in order to eliminate SARS-CoV-2.

## Data Availability

All data produced in the present study are available upon reasonable request to the authors.

## Author contributions

DJ, MaB, AD and NSM conceived the study. DJ, MaB, TRW, SM, AZ, UR, AD and NSM planned experiments. AD and NSM supervised the study. DJ, MaB, PDK, TRW, SM, TG, JoG, PM, JeG and BT performed experiments. CH, YTP, JH, RF, AK, AN, YM, SS, JSW, KA, GU, MM, TB, KSLSB, HH, SG, MiB, HR, JR, CE, ARF, MH, BK, MS and GK collected samples or organised their collection. KSL and NSM obtained funding. MaB and AD performed data analysis and generated the figures. AD wrote the first draft of the manuscript. All authors approved the manuscript prior to submission.

## Acknowledgements

We would like to thank other members of the Multiplex Immunoassays, Biological Development Center and Bioanalytics groups at the NMI for their support on this project. We would like to thank Ann Kathrin Horlacher and Mareike Walenta for assistance with sample processing and patient material storage. We Ulrike Schmidt, Iris Schaefer, Richard Schaad and Hannah Zug for technical support. We thank Katharina Kienzle, Hartmut Mahrhofer, Hardy Richter, Stefanie Döbele for help with sample collection. We thank Andrea Evers-Bischoff, Andrea Bevot and the CPCS at the University Hospital Tuebingen for organizational support in conducting the study.

We would like to thank all those involved in the organisation of the MuSPAD and TuSeRe sample collection. This work was financially supported by the State Ministry of Baden-Württemberg for Economic Affairs, Labour and Housing Construction (grant numbers FKZ 3-4332.62-NMI-67 and FKZ 3-4332.62-NMI-68), the Initiative and Networking Fund of the Helmholtz Association of German Research Centres (grant number SO-96), the EU Horizon 2020 research and innovation program (grant agreement number 101003480 - CORESMA). The funders had no role in study design, data collection and analysis, decision to publish, or preparation of the manuscript.

