## Supplementary Information for "Antibody binding and ACE2 binding inhibition is significantly reduced for the Omicron variant compared to all other variants of concern"

21 – TWINCORE, Centre for Experimental and Clinical Infection Research, a joint venture of Hannover Medical School and the Helmholtz Centre for Infection Research, Hannover, Germany

* indicates shared first authorship

§ indicates shared last authorship

### indicates corresponding author

**Corresponding author Information**

Alex Dulovic –, +49 (0)7121 51530 580

Nicole Schneiderhan-Marra –, +49 (0)7121 51530 815

#### Methods

##### Sample Cohort

Serum samples used in this study were originally collected for use in several other studies (1-4). The sample set was designed to include a broad representation of infected samples from the different waves of SARS-CoV-2 within Germany, as well as vaccinated samples. We separated our cohort into 4 groups: vaccinated samples, convalescent samples, infected and later vaccinated samples and pre-pandemic samples as controls.

Vaccinated study participants received either two homologous doses of AZD1222, BNT162b2 or mRNA-1273, two heterologous doses of AZD1222 and BNT162b2 or AZD1222 and mRNA-1273, or three doses of BNT162b2. To examine how antibody dynamics changed over time, we included samples from both one to two months post-second dose and four to six months post-second dose. We also included samples from vaccinated individuals who had a previous PCR-confirmed infection and then received a single dose of BNT162b2 as per national guidelines. Convalescent study participants were collected approximately three months post-positive PCR. To assess differences between variants of SARS-CoV-2, we collected samples from those with a WT, alpha or delta infection. WT samples came from both adults and children. To be considered a WT sample, the infection had to occur during the first wave of the SARS-CoV-2 in Germany (spring-summer 2020). Alpha samples were confirmed by PCR sequencing and were collected between January to May 2021. Delta samples were either confirmed by PCR sequencing, or where collected during a time period when all infections in Germany were considered to be delta (September-November 2021). To represent naïve samples, negative pre-pandemic samples were obtained from Central BioHub. An overview of the characteristics for each cohort sub-group can be found as Table 1.

**Ethical Oversight**

Informed written consent was obtained from all study participants. Ethical approval and oversight for the samples used in this study was provided by the following ethics committees: the Ethics Committee of the University Hospital Tuebingen (293/2020BO2, 764/2020/BO2 (amended 6.12.21), B312/2020BO1 (amended 02.06.21), 556/2021BO1), the Ethics Committee of the University of Tuebingen (179/2020/BO2, 188/2020A) and the Ethics Committee of Hannover Medical School (9086_BO_S_2020).

**Mass Spectrometry of Omicron receptor binding domain (RBD)**

The RBD omicron protein samples (5 µg) were N-deglycosylated using a PNGaseF reducing kit (Rapid PNGaseF reducing kit, New England Biolabs, Frankfurt am Main, Germany) by adding ¼ of the volume of reducing buffer included in the kit and denaturation and reduction for 5 minutes at 80 °C. Subsequently, 0.125 µL PNGaseF enzyme preparation, included in the kit, were added, deglycosylation was performed for 10 minutes at 50 °C. Prior to Liquid chromatography-mass spectrometry (LC-MS) analysis, the samples were diluted 1:3 with HisNaCl buffer (20 mmM His 140 mM NaCl, pH 6.0) and analyzed by liquid chromatography (HPLC) coupled to electrospray ionization (ESI) quadrupole time-of-flight (QTOF) MS. Samples (0.4 µg per injection) was desalted using reversed phase chromatography on a Dionex U3000 RSLC system (Thermo Scientific, Dreieich, Germany) using a Acquity BEH300 C4 column (1mm x 50mm, Waters, Eschborn, Germany) at 75°C and 150 µl/min flow rate applying a 11-min linear gradient with varying slopes. In detail, the gradient steps were applied as follows (min/% Eluent B): 0/5, 0.4/5, 2.55/30, 7/50,

7.5/99, 8/5, 8.75/99, 9.5/5, 10/99, 10.25/5 and 11/5. Eluent B was acetonitrile with 0.1% formic acid, and solvent A was water with 0.1% formic acid. To avoid contamination of the mass spectrometer with buffer salts, the HPLC eluate was directed into waste for the first 2 min. Continuous MS analysis was performed using a QTOF mass spectrometer (Maxis UHR-TOF; Bruker, Bremen, Germany) with an ESI source operating in positive ion mode. Spectra were taken in the mass range of 600–2000 m/z. External calibration was applied by infusion of tune mix via a syringe pump during a short time segment at the beginning of the run. Raw MS data were lock-mass corrected (at m/z 1221.9906) and further processed using Data Analysis 5.3 and MaxEnt Deconvolution software tools (Bruker).

**Antigen Immobilisation on beads**

SARS-CoV-2 wild-type Spike, RBD, S1 domain, S2 domain and Nucleocapsid were immobilised on magnetic MagPlex beads (Luminex) by EDC-sNHS coupling as previously described(5).RBDs from variants of concern and the Omicron Spike protein were immobilised on magnetic MagPlex beads (Luminex) by Anteo coupling (AMG Activation Kit for Multiplex Microspheres, #A-LMPAKMM-400, Anteo Technologies) as previously described(2). Following coupling, beads were stored at 4°C. Prior to experimentation, beads were then combined into a 25x Bead Mix and stored at 4°C until used. The antigens used in these experiments can be found as Table 2.

**MULTICOV-AB**

MULTICOV-AB, a previously published multiplex immunoassay was performed as described(5). A full list of antigens included within the assay are listed in Table 2. Samples were randomly allocated to plates to ensure that at least sample of every sample group was included on each plate. Briefly, samples were thawed at room temperature, vortexed and then diluted 1:200 in assay buffer before being mixed 1:1 with 1x Bead Mix in a 96-well plate (final dilution 1:400). Samples were then incubated in darkness on a Thermomixer (20°C, 750 rpm, 2 hours) before being washed three times to remove unbound antibodies. To ensure retention of beads, a magnetic plate washer was used. To detect bound IgG, 3 µg/mL RPE-goat anti-human IgG was added to each well and then incubated for a further 45 mins on a Thermomixer. After another washing step, beads were resuspended in 100 µL of wash buffer, shaken for 3 mins at 1000 rpm, and then measured once on a FLEXMAP3D instrument (No Timeout, Gate 7500-15000, Reporter Gain Standard PMT, 50 events). 3 Quality control samples were included in duplicate on each plate. All samples were measured twice in two independent experiments. No sample failed QC. Raw median fluorescence intensity (MFI) values were normalized to a QC sample for all antigens as per(6) .

**RBDCoV-ACE2**

RBDCoV-ACE2, a previously published multiplex ACE2 inhibition assay(2), analyzes neutralizing antibody activity through ACE2 binding inhibition. A full list of antigens included in this assay can be found as Table 2. Briefly, 1:25 diluted samples from MULTICOV-AB, were further diluted to 1:200 in ACE2 buffer(2), which contains 300 ng/mL biotinylated ACE2. Samples were then mixed 1:1 with 1x VOC bead mix in 96 well plates and incubated for 2 hours in darkness on a thermomixer (750 rpm, 20°C). Following this initial incubation, samples were washed to remove unbound ACE2 using an automated magnetic plate washer. Bound ACE2 was detected by adding 2 µg/mL RPE-labelled streptavidin and incubating for a further 45 mins. After washing to remove unbound fluorophores, beads were resuspended in 100 µL washing buffer and shaken for 3 mins at 1000 rpm. Plates were measured once on a FLEXMAP3D instrument (No Timeout, Gate 7500-15000, Reporter Gain Standard PMT, 50 events). As controls, 3 wells with 150 ng/mL ACE2, 2 blank wells and 3 wells with a QC sample were included. ACE2 binding inhibition was calculated as a percentage, with 100% indicating maximum ACE2 binding inhibition and 0% indicating no ACE2 binding inhibition. Samples with an ACE2 binding inhibition less than 20% are classified as non-responders(2).

**Biolayer Interferometry (BLI)**

Purified $RBD_{wt}$, $RBD_{\delta}$ and $RBD_{o}$ were biotinylated with Sulfo-NHS-LC-LC-Biotin (Thermo Fisher Scientific) in 5 molar excess at ambient temperature for 30 min. Excess of biotin was removed by size exclusion chromatography using Zeba$^{TM}$ Spin Desalting Columns 7K MWCO 0.5 ml (Thermo Fisher Scientific) according to manufacturer's protocol. Analysis of binding kinetics of RBD specific antibodies in serum samples were performed using the Octet RED96e system (Sartorius) as per the manufacturer's recommendations. In brief, 5 µg/ ml of each biotinylated RBD diluted in Octet buffer (PBS, 0.1% BSA, 0.02% Tween20) was immobilized on streptavidin coated biosensor tips (SA, Sartorius) for 20 s. In the association step, serum samples at a 1:100 dilution were reacted for 720 s followed by dissociation in Octet buffer for 1200 s. Every run was normalized to a healthy control sample lacking RBD specific antibodies and for each sample technical duplicates (n = 2) were performed. Data were analyzed using the Octet Data Analysis HT 12.0 software applying the 1:1 fitting model for the dissociation step. The binding profile response of each sample is illustrated as the mean wavelength shift in nm. Binding kinetics for ACE2 were performed by immobilizing 5 µg/ ml of each biotinylated RBD diluted in Octet buffer on streptavidin coated biosensor tips (SA, Sartorius) for 20 s. Dilution series ranging from 50 to 6.25 nM of ACE2 (Sino Biological) were applied for 300 s and one reference was included per run, followed by a dissociation step in Octet buffer (480s). For affinity determination, the 1:1 global fit of the Data Analysis HT 12.0 software was used.

**Data Analysis**

Data was collated and matched to metadata in Excel 2016. Data visualisation was done in RStudio (Version 1.2.5001 running R version 3.6.1). Additional packages “gplots” and “beeswarm” were used for specific displays. The “lm” function of R’s “stats” library was used for linear regression analyzes. Correlation analyzes were performed using the “cor” function of R’s “stats” library. The “wilcox.test” function from R’s “stats” library was used to perform Mann-Whitney-U Tests (two-sided) in order to estimate significance of observed differences between different groups. Graphs were exported from RStudio and further edited in Inkscape (Version 0.92.4) to generate final figures. Biolayer inferometry graphical representation was prepared using GraphPad Prism Software (Version 9.0.0).

**Supplementary Figures**

Supplementary Figure 1: **Binding kinetics of RBD specific antibodies from serum samples of vaccinated and convalescent individuals.**

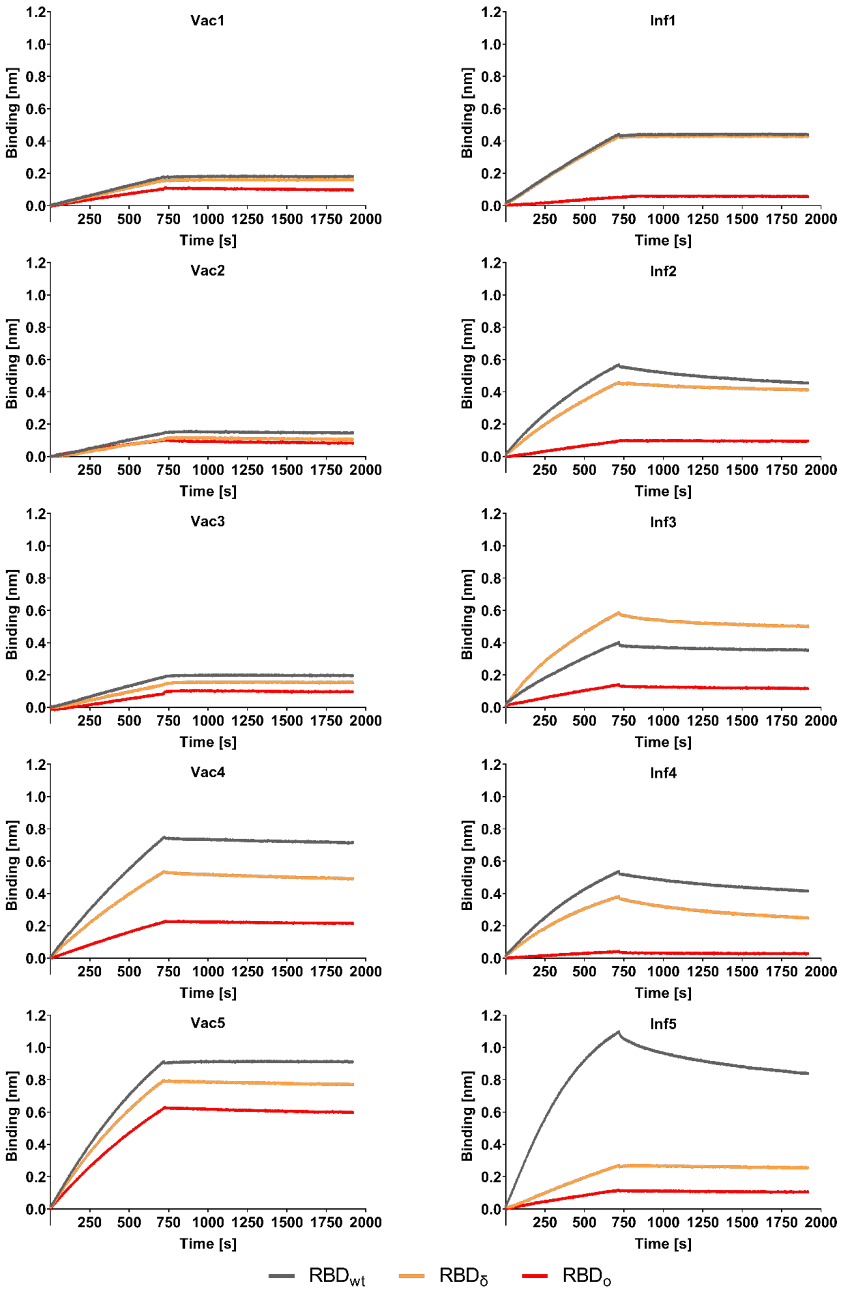


Biotinylated RBDwt, RBDδ and RBDo were immobilized on streptavidin biosensor tips and binding kinetics of serum samples from vaccinated (n = 5, Vac) and convalescent (n =5, Inf) individuals were analyzed using BLI. All sensograms are illustrated as mean of technical duplicates (n = 2).

Supplementary Figure 2: **Binding kinetics of ACE2 to $RBD_{wt}$, $RBD_{\delta}$ and $RBD_{o}$ using BLI.**

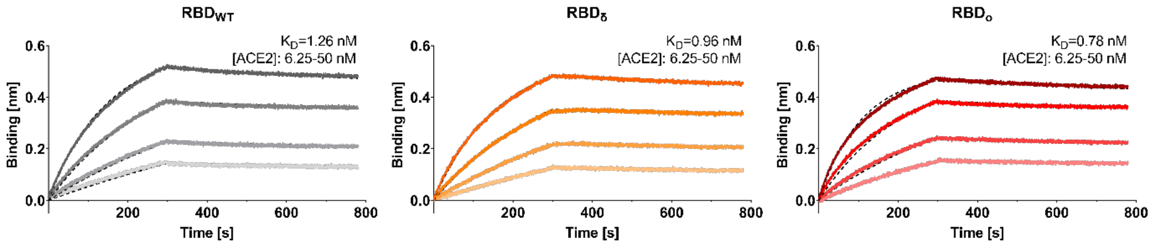


| | $K_D$ [nM] | $k_{on}$ [$10^5$ $M^{-1}$ $s^{-1}$] | $k_{off}$ [$10^{-4}$ $s^{-1}$] |
|---|---|---|---|
| **$RBD_{wt}$** | 1.26 ± 0.01 | 1.37 ± 0.00 | 1.72 ± 0.01 |
| **$RBD_{\delta}$** | 0.96 ± 0.01 | 1.24 ± 0.00 | 1.19 ± 0.01 |
| **$RBD_{o}$** | 0.78 ± 0.01 | 1.85 ± 0.00 | 1.45 ± 0.01 |

For biolayer interferometry (BLI)-based affinity measurements, biotinylated $RBD_{wt}$, $RBD_{\delta}$ and $RBD_{o}$ were immobilized on streptavidin biosensors. Kinetic measurements were performed using four concentrations of purified ACE2 ranging from 6.25 nM to 50 nM (illustrated with gradually lighter shades). The table summarizes affinities ($K_D$), association ($k_{on}$), and dissociation constants ($k_{off}$) of ACE2 determined for the different RBD variants.

Supplementary Figure 3: **ACE2 binding inhibition for two-dose BNT162b2, 1-2 months post-administration of the second dose**.

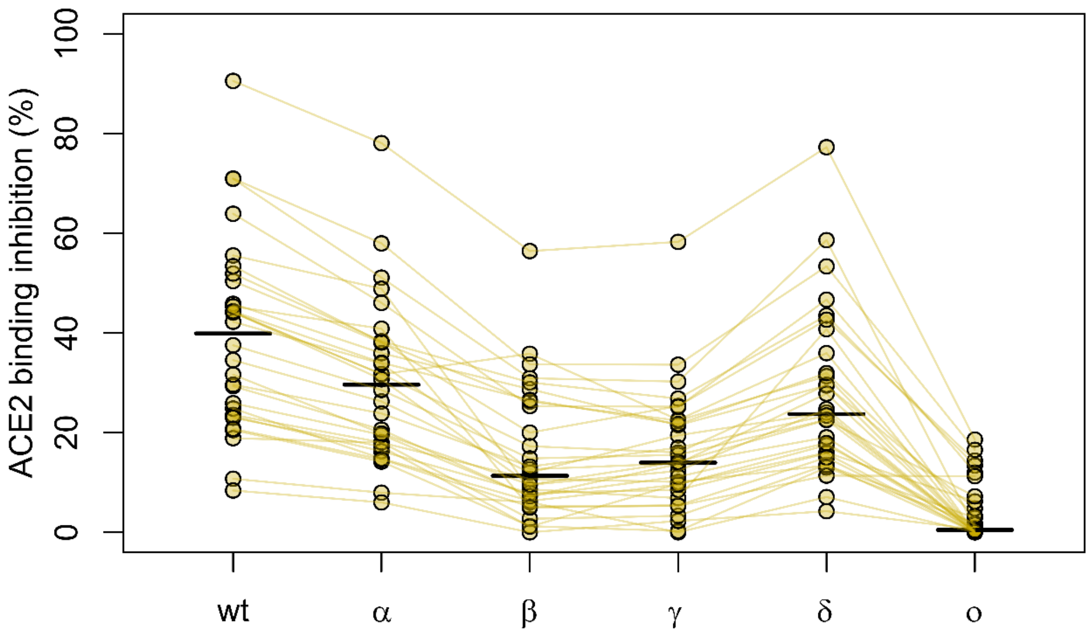


Comparative ACE2 binding inhibition for all variants of concern and wild-type as calculated with RBDCoV-ACE2. Lines indicate paired samples. The equivalent figures for 5-6 months post-second dose and following the 3rd dose can be found as Supplementary Figure 5 and Figure 3c respectively.

Supplementary Figure 4: **ACE2 binding inhibition for two-dose BNT162b2, 5-6 months post-administration of the second dose.**

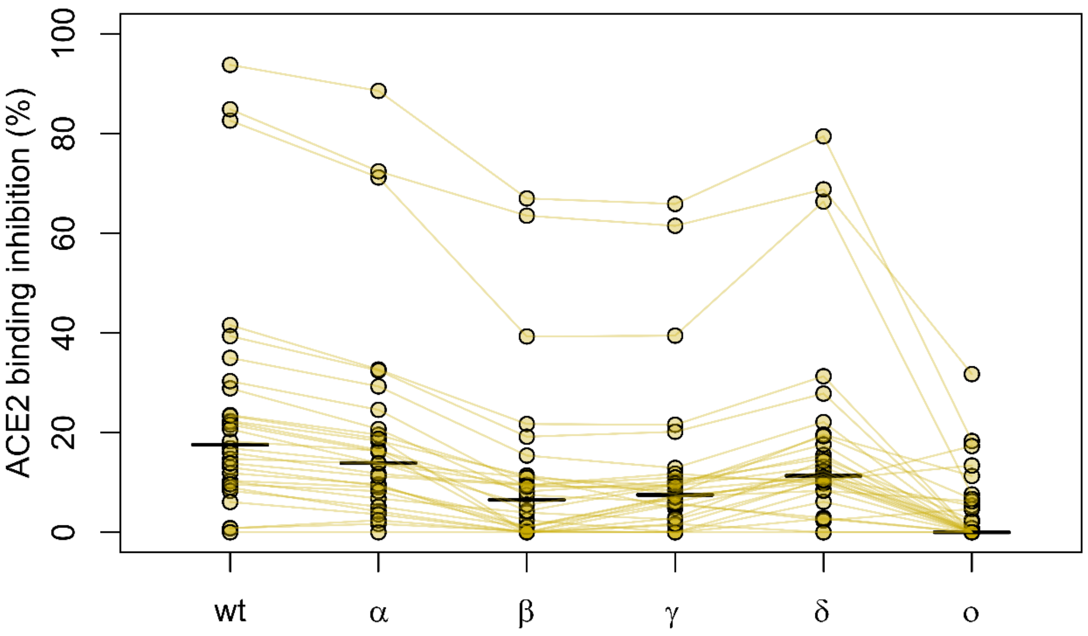

Supplementary Table 1: **Median fold reduction in binding from wild-type to respective variant**

| Group | Vaccinated | Infected |
|---|---|---|
| WT | 1.00 | 1 |
| alpha | 1.13 | 1.11 |
| beta | 2.59 | 3.16 |
| gamma | 1.67 | 1.99 |
| delta | 1.24 | 1.30 |
| omicron | 4.51 | 8.65 |
| lambda | 1.64 | 1.96 |
| mu | 2.90 | 3.45 |

Supplementary Table 2: **Median fold change reduction from respective variant to Omicron**

| Group | Vaccinated | Infected |
|---|---|---|
| WT | 4.51 | 8.65 |
| alpha | 3.96 | 7.41 |
| beta | 1.74 | 2.36 |
| gamma | 2.67 | 4.04 |
| delta | 3.55 | 6.34 |
| omicron | 1.00 | 1.00 |
| lambda | 2.67 | 4.03 |
| mu | 1.53 | 2.15 |

Supplementary Table 3: **Median ACE2 binding inhibition of the respective variant**

| Group | Vaccinated | Infected |
|---|---|---|
| WT | 25.62 | 33.26 |
| alpha | 19.69 | 33.56 |
| beta | 8.92 | 8.83 |
| gamma | 10.00 | 8.28 |
| delta | 18.01 | 19.78 |
| omicron | 0.00 | 1.91 |
| lambda | 18.43 | 15.00 |
| mu | 7.95 | 6.78 |

Supplementary Table 4: **Summary of the binding response and dissociation constant ($k_{off}$) of the individual serum samples.**

| | Response [nm] | | | Dissociation constant $k_{off}$ [$10^{-5}$ $s^{-1}$] | | |
|---|---|---|---|---|---|---|
| | **$RBD_{wt}$** | **$RBD_{\delta}$** | **$RBD_{o}$** | **$RBD_{wt}$** | **$RBD_{\delta}$** | **$RBD_{o}$** |
| **Vac1** | 0.175 ± 0.008 | 0.154 ± 0.018 | 0.102 ± 0.008 | 0.010 ± 0.000 | 0.010 ± 0.000 | 8.620 ± 1.287 |
| **Vac2** | 0.143 ± 0.005 | 0.104 ± 0.005 | 0.098 ± 0.016 | 3.895 ± 0.148 | 7.880 ± 1.824 | 14.400 ± 4.808 |
| **Vac3** | 0.186 ± 0.011 | 0.139 ± 0.002 | 0.082 ± 0.009 | 0.010 ± 0.000 | 0.010 ± 0.000 | 5.060 ± 2.065 |
| **Vac4** | 0.741 ± 0.034 | 0.531 ± 0.020 | 0.222 ± 0.015 | 3.080 ± 0.170 | 6.050 ± 0.269 | 4.520 ± 0.877 |
| **Vac5** | 0.908 ± 0.019 | 0.791 ± 0.045 | 0.618 ± 0.030 | 0.010 ± 0.000 | 2.200 ± 0.099 | 3.830 ± 0.552 |
| **Inf1** | 0.439 ± 0.009 | 0.425 ± 0.072 | 0.052 ± 0.002 | 0.010 ± 0.000 | 0.181 ± 0.242 | 0.136 ± 0.178 |
| **Inf2** | 0.565 ± 0.015 | 0.456 ± 0.011 | 0.095 ± 0.007 | 16.700 ± 0.141 | 7.630 ± 0.184 | 3.525 ± 0.290 |
| **Inf3** | 0.399 ± 0.072 | 0.581 ± 0.099 | 0.139 ± 0.010 | 5.635 ± 0.502 | 10.050 ± 0.071 | 8.655 ± 1.775 |
| **Inf4** | 0.534 ± 0.019 | 0.380 ± 0.039 | 0.039 ± 0.005 | 18.750 ± 0.212 | 32.350 ± 0.354 | 17.450 ± 2.758 |
| **Inf5** | 1.094 ± 0.025 | 0.267 ± 0.010 | 0.115 ± 0.018 | 18.800 ± 0.283 | 4.315 ± 0.516 | 6.185 ± 2.609 |

Binding kinetics of serum samples from (n = 5, Vac) and convalescent (n =5, Inf) individuals to the different RBD variants were analyzed using BLI. Binding response and dissociation constant ($k_{off}$) determined by the 1:1 fitting model of the individual serum samples were summarized as mean ± SD.
